# Urogenital schistosomiasis in women of reproductive age in Kilifi County, Kenya

**DOI:** 10.64898/2026.03.24.26349244

**Authors:** Hellen Wambui Kariuki, Sophia Mogere Nyasore, Felistas Muthini, Peter Waweru Mwangi, Jackson Muinde Mwandi, Patrick Makazi, Marianne Wanjiru Mureithi, Wallace Bulimo, Tim Joash Lugera Wango, Eric Wanjala, Lyle Mckinnon, Kariuki Njaanake

## Abstract

**Background:** Urogenital schistosomiasis (UGS), caused by Schistosoma haematobium (S. haematobium,) disproportionately affects women in sub-Saharan Africa and can lead to haematuria, anaemia, and urinary tract morbidity. Data on the prevalence in women of reproductive age remains limited in contrast to infection among school-aged children in Kenya. This study assessed the prevalence of UGS and its socioeconomic determinants among women in Kilifi County, Kenya.

**Methods:** Urine samples (20–50 mL) were collected from each participant over three consecutive days. Day-one samples were tested for haematuria, proteinuria, nitrites, leukocytes, and pregnancy using dipsticks. On the other hand 10 mL of urine was examined for *S. haematobium* eggs via urine filtration all three days.

**Results:** A total of 599 women aged 15–50 years were enrolled, with complete data available for 336. The mean age was 33 years; 57.7% were <35 years. Most participants were from rural Magarini Sub-county (63%) and engaged in crop farming (62.5%). Primary education was the highest level attained by 59.8% of participants. Frequent contact with stagnant water was reported by 92%. The overall prevalence of *S. haematobium* infection was 13.7% (95% CI: 10.2–17.8), higher in Magarini (14.9%) than in Rabai (12.0%), though not statistically significant. Younger age, primary education, and frequent water contact were associated with higher infection rates; however, after adjustment for covariates, haematuria showed the strongest independent association with infection. Women with haematuria were 25.2 times more likely to be infected (OR: 25.24, 95% CI: 7.07–82.63, p < 0.001); multivariate analysis confirmed haematuria as the sole significant predictor (OR: 20.83, 95% CI: 5.45–79.57, p < 0.001).

**Conclusion:** Urogenital schistosomiasis prevalence among women in Kilifi County is substantial, with variation between sub-counties. Haematuria demonstrated the strongest independent association with infection and may serve as a simple, non-invasive diagnostic marker. These findings underscore the pressing need for the integration of UGS screening into the reproductive health services and targeted interventions.

**Authors Summary:** Urogenital schistosomiasis, caused by the parasitic worm Schistosoma haematobium, is a neglected tropical disease and remains a major public health problem in sub-Saharan Africa. Although control programmes in Kenya primarily target school-aged children, women of reproductive age are frequently exposed through daily water contact and may develop chronic urinary and reproductive health complications. However, data on the infection burden among adult women are limited. In this study, we assessed the prevalence of urogenital schistosomiasis and associated risk factors among women aged 15–50 years in Kilifi County, Kenya. Urine samples were collected over three consecutive days and examined for parasite eggs and indicators of urinary tract disease. We found that urogenital schistosomiasis affected more than one in ten women in the rural sub-counties where the study was conducted. Blood in urine (haematuria) was strongly associated with infection and remained the most reliable predictor after accounting for other social and behavioural factors. These findings demonstrate that urogenital schistosomiasis is an under-recognised health issue among women and highlight the potential value of simple urine-based screening tools. Integrating urogenital schistosomiasis screening into existing reproductive health services could improve early detection and contribute to more inclusive disease control strategies.

## Introduction

Urogenital schistosomiasis (UGS) is a neglected tropical disease caused by the trematode *Schistosoma haematobium* (1). In women, UGS presents with symptoms such as dysuria, haematuria, unusual vaginal discharge, genital pain, vaginal bleeding, and genital lesions (2,3). Transmission of urogenital schistosomiasis depends on *Bulinus* spp. snails, which serve as intermediate hosts by releasing cercariae into freshwater. Humans become infected when the cercariae penetrate the skin during contact with contaminated water (4,5). Women are particularly at risk due to frequent exposure while performing domestic chores such as fetching water or washing clothes (6). If left untreated, urogenital schistosomiasis can lead to serious complications, including bladder cancer, infertility, cervical cancer, adverse pregnancy outcomes, preterm labour, and increased susceptibility to HIV infection (7–10). The World Health Organisation recommends annual preventive chemotherapy via mass drug administration campaign with praziquantel (PZQ) in endemic regions (2).

With an estimated prevalence of 17.5% among women, urinary schistosomiasis (UGS) remains a significant but often neglected public health concern in sub-Saharan Africa (11). A study conducted in the Kileo area of Tanzania reported a prevalence of 2.3% among women of reproductive age in the (6) while in Kenya, the coastal counties of Kwale, Kilifi, Lamu, Mombasa, and Tana River are the most affected, with prevalence among women of reproductive age reaching up to 22.3% in Kilifi County (12–14). The effective management of UGS among women remains challenging. Awareness and diagnostic capacity among healthcare workers are limited, leading to frequent misdiagnosis and underreporting (9,15,16). For instance, a study in Zanzibar found that many healthcare workers believed UGS primarily affects boys and men, as women rarely present with haematuria (17). Additionally, stigma and embarrassment associated with genital or urinary symptoms, often mistaken for sexually transmitted infections, discourage many girls and women from seeking timely treatment (18,19).

Although previous studies have documented the prevalence of UGS in coastal Kenya, there is limited research that examines the factors that sustain transmission among women. Most African studies on UGS have primarily focused on school-aged children, with little attention given to equally vulnerable women (14). This study, therefore, aimed to address this gap by determining the prevalence and key risk factors of UGS among women of reproductive age in Kilifi County, Kenya. The research was conducted specifically in Rabai and Magarini Sub-counties, which are recognised hotspots for *Schistosoma haematobium* transmission due to their proximity to freshwater bodies and frequent community water contact activities. The study focused on sociodemographic characteristics, clinical outcomes based on urinalysis, and water contact behaviours, including sources of water for domestic use and frequency of exposure to stagnant water.

## Methodology

### Study design

This cross-sectional study was conducted in September, October and November 2022. Women of reproductive age between 15 and 50 years were enrolled in the study after obtaining informed consent randomly recruited in Rabai and Magarini sub-counties in Kilifi County of coastal Kenya. Urine specimens (20–50 mL) were collected from each participant between 10:00 a.m. and 2:00 p.m. on three consecutive days. For each specimen, 10 mL was processed and examined for *Schistosoma haematobium* eggs using standard parasitological techniques.

### Study Area

This study was conducted in Kilifi County (population of 1,453,787), located along the Kenyan coast, bordering Mombasa to the south, Kwale to the southwest, Tana River to the north, and Taita Taveta to the west. The county has a warm tropical climate, with temperatures ranging from 21°C to 32°C (20). Most residents are subsistence farmers, fisherfolk, and informal traders. The study specifically focused on the Rabai and Magarini sub-counties, which are recognised hotspots for *Schistosoma haematobium* transmission due to frequent human–water contact and the presence of *Bulinus* snail habitats in stagnant and slow-moving freshwater sources (21).

The residents have limited access to piped water, which results in widespread reliance on rivers, dams, and seasonal swamps for domestic and agricultural activities, which sustains ongoing transmission of urinary schistosomiasis despite intermittent piped water availability in some areas (**Figure 1**).

**Figure 1:**
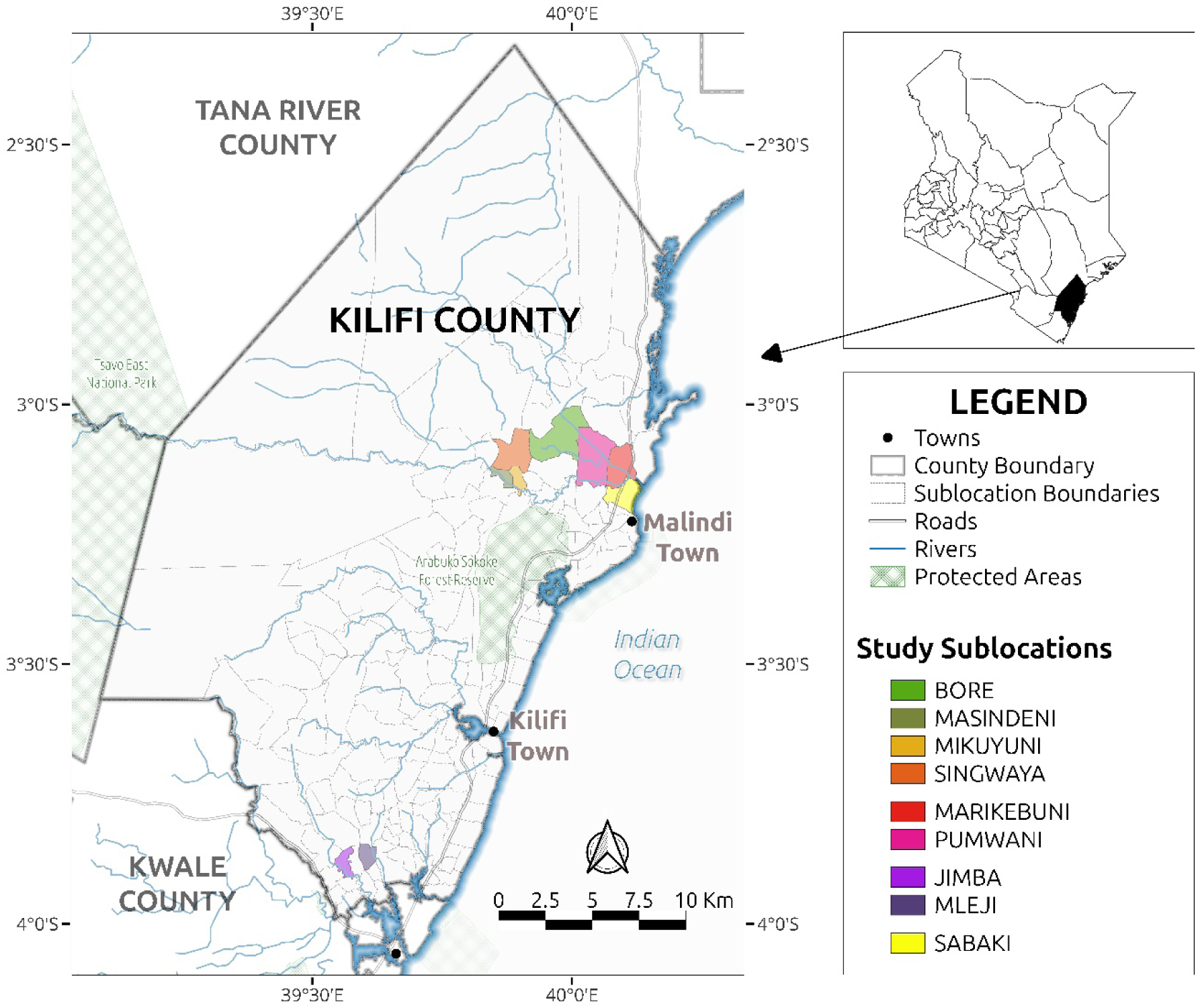
Map of Kilifi County, Kenya, showing the study area. The map illustrates the geographical context of the study, highlighting sub-counties and sub-locations within Kilifi County. Key physical and administrative features, including major rivers, roads, and county boundaries, are also depicted to provide spatial orientation and reference points relevant to the study.

### Sampling technique

Rabai and Magarini sub-counties were purposively selected as *S. haematobium* hot spots based on data from the Kilifi Neglected Tropical Diseases Department, which had previously reported a high prevalence of urinary schistosomiasis among school-going children in these areas (22). In Magarini, participants were recruited from Pumwani, Marikebuni, Bore in Magarini ward, Singwanya, Masindeni and Mikuyuni sub-locations in Garashi and Sabaki wards, while in Rabai, recruitment was performed in Jimba and Mleji sub-locations within Ruruma Ward. A comprehensive list of eligible households was compiled in each village from which 150 participants were randomly selected per sub-county. Only one participant was enrolled per household. When more than one eligible individual was present, a participant was chosen using a simple random (lottery) method to prevent intra-household clustering.

### Sample size estimation

Due to a lack of data on the prevalence of UGS among adults in the study area, we used a conservative prevalence range of 10 – 20%, based on prior adult-focused studies (2.6 – 18%) from coastal Kenya and a global meta-analysis estimate of 17.5% among women of reproductive age. The required sample per sub-county was calculated using the single-proportion formula *n₀ = Z²p(1−p)/d²* with 95% confidence (Z = 1.96) (23). Scenarios with p = 0.10 – 0.20 and precision d = 0.05 yielded base sample sizes of 138 – 246 per stratum, which increased to approximately 200–355 after adjusting for clustering (design effect, DEFF = 1.3) and a 10% non-response rate. Due to logistical constraints in Phase I, we selected d = 0.07 at p = 0.15, producing a target of about 144 participants per sub-county after adjustment for DEFF and non-response. Consequently, we aimed for around 150 participants in each sub-county (a total ≈ 300). The calculated sample size provided adequate precision for prevalence estimation and sufficient power for multivariable analysis. Participants were selected through proportional allocation across villages within each sub-county based on the number of women of reproductive age. Eligible households were visited systematically using a pre-determined sampling interval, and one eligible woman per household was randomly selected.

### Questionnaire administration

A questionnaire on risk factors for *S. haematobium* infection, such as economic activities, water-related activities, education level, and residential ward, was administered to each participant at the time of urogenital sampling (shown in supplementary material).

### Urine collection and microscopy

A 10 ml urine sample from each participant was examined for *S. haematobium* eggs for three consecutive days using the filtration technique. A 12 μm pore polycarbonate filter was used and examined under a microscope at X40 magnification (19). *S. haematobium* eggs were counted, and an average of the three-day count was calculated. Infections were classified into negative for no eggs, light (1–49 eggs/ 10 ml urine) or heavy (≥50 eggs/10 ml urine) (24). On day one, a part of the urine sample was also used for point-of-care testing, including urinalysis (hematuria, proteinuria, nitrites, and leukocytes) using Uristix*®* reagent strips and pregnancy testing with the ACCURATE^®^ (one-step pregnancy test strip). The remaining 40 ml of urine was stored in cryovials for future analysis and transported to the Department of Medical Microbiology and Immunology laboratory, University of Nairobi.

### Data entry, cleaning, and analysis

Data were entered in a Microsoft Excel spreadsheet (Microsoft^®^Corp., Redmond, WA, USA) and analysed using IBM SPSS Statistics for Windows, Version 26 (Released 2018; IBM Corp., Armonk, New York, USA). Pearson’s Chi-squared test was used to assess the association between categorical variables. A univariate model was initially used to screen variables for their association with *S. haematobium* infection. Binomial logistic regression was used to assess the combined effects of multiple variables. Factors with *p*-values less than 0.25 were included in a multivariate logistic regression model to identify significant predictors of infection while controlling for potential confounders (25). In all tests, *p*-values less than 0.05 were regarded as statistically significant.

### Ethical approval and considerations

Ethical clearance was obtained from the Kenyatta National Hospital–University of Nairobi Ethics and Research Committee (KNH-ERC) (approval number P334/05/2018) and the National Commission for Science, Technology and Innovation (NACOSTI) (reference number NACOSTI/P/22/1769). Permission was also obtained from the Kilifi County Health Research Committee, Kilifi County Government, through the County Commissioner and the Kilifi County Education Office. The study included participants aged 15–17 years, classified as emancipated minors under Kenyan ethical guidelines. In accordance with national regulations and with approval from the KNH-ERC, these participants provided written informed consent independently, and parental or guardian consent was therefore not required. All participants who tested positive for *S. haematobium* eggs in urine were treated with praziquantel (Bayer AG, Germany) at a dose of 40 mg/kg, in line with national treatment guidelines.

## Results

### Sociodemographic characteristics of the study population

Risk factor analysis was performed for participants with complete datasets (N=336). The analysis focused on identifying socio-demographic and behavioural factors associated with urinary schistosomiasis infection. A total of 336 women of reproductive age participated in the study, drawn from Rabai and Magarini sub-counties of Kilifi County. Of these, 142 (42.3%) were from Rabai and 194 (57.7%) from Magarini. In Rabai, participants were enrolled from Ruruma Ward, specifically Jimba and Mleji sub-locations, while in Magarini, participants were drawn from Magarini, Garashi, and Sabaki wards. The detailed distribution of participants by sub-county, ward, and sub-location is shown in **Table 1**. Most participants (62.5%) were engaged in farming as their primary economic activity, followed by business (17.0%) and domestic work (16.6%). Only a small proportion (3.9%) were casual labourers. The majority (93.2%) had either no formal education or only primary-level education, while 6.8% had completed secondary education.

**Table 1.**
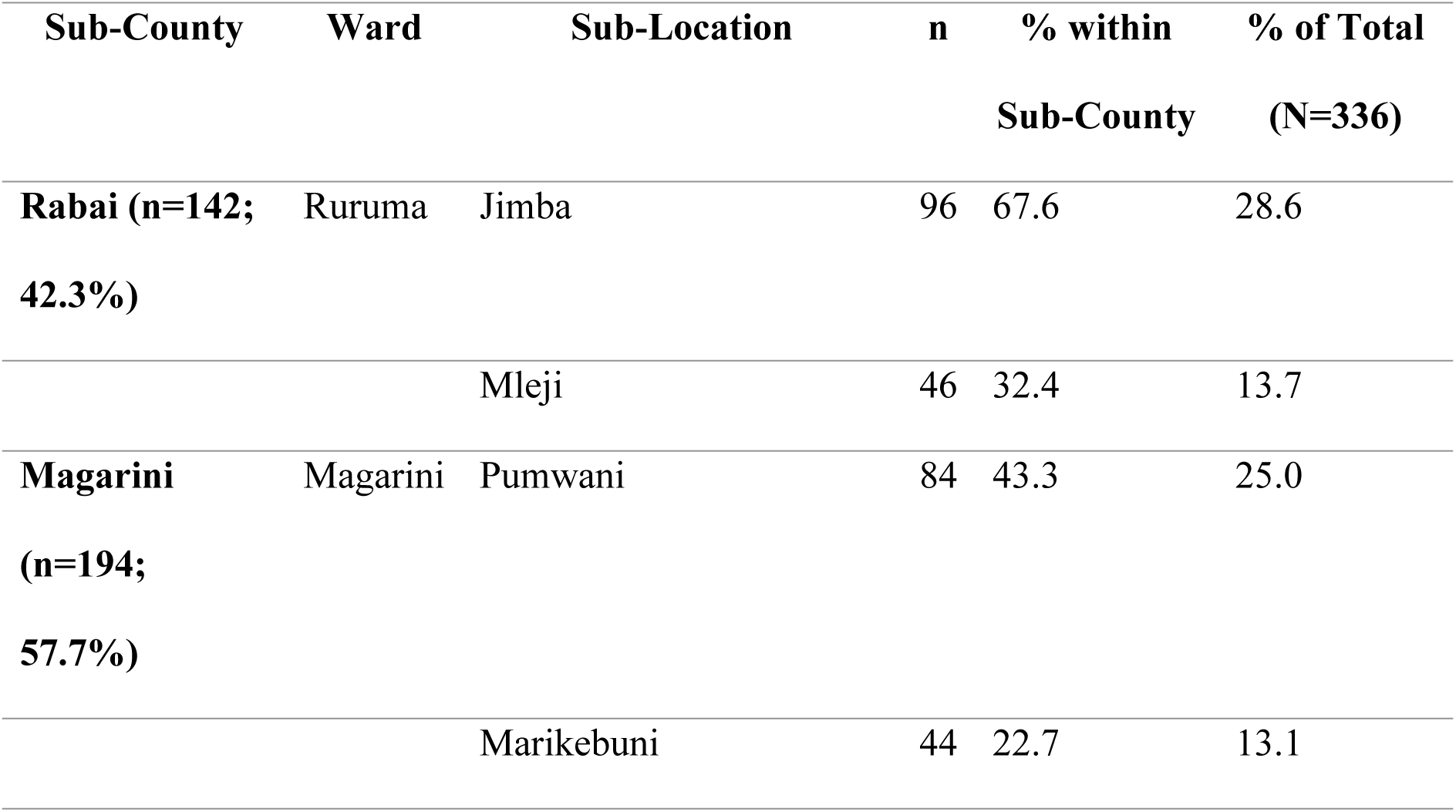
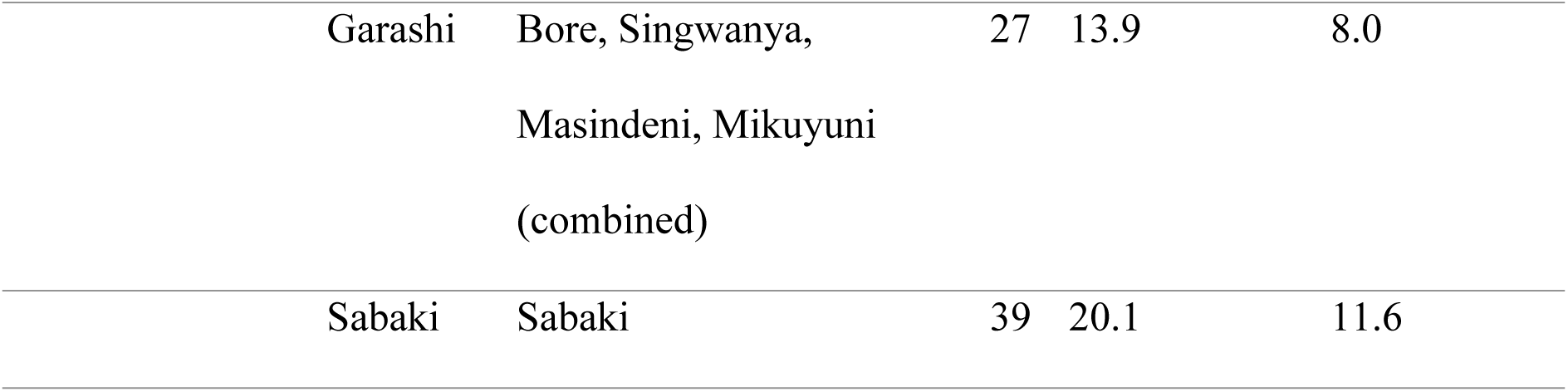
Distribution of participants by Sub-County, Ward, and Sub-Location.

### Water Contact and Exposure Patterns

Rivers were the most common source of domestic water (48.2%), followed by ponds (20.2%) and piped water (13.4%). A small proportion of participants reported using mixed water sources, including combinations of piped water, wells, and rivers. Overall, 92% of the participants reported contact with stagnant water. The most common activities associated with such contact included washing or fetching water (81.9%), bathing (64.6%), and walking through stagnant water (11.0%). Multiple forms of contact were also reported, with some women engaging in two or more of these activities. Most participants (69.0%) reported exposure to stagnant water more than twice a week, while 16.4% experienced seasonal exposure and 6.5% had exposure less than twice a week. A previous history of urinary schistosomiasis infection was reported by 17% of the participants, and of these, 84.2% had sought treatment. The socio-demographic, water contact, and exposure characteristics are summarised in **Table 2**.

**Table 2.** Socio-Demographic and water contact characteristics of participants (N=336)

| Variable | Characteristics | n | % |
| --- | --- | --- | --- |
| Occupation | Farming | 210 | 62.5 |
|  | Business | 57 | 17.0 |
|  | Housewife | 56 | 16.6 |
|  | Casual labourer | 13 | 3.9 |
| **Education level** | No formal/Primary | 313 | 93.2 |
|  | Secondary | 23 | 6.8 |
| **Main source of domestic water** | River | 162 | 48.2 |
|  | Pond | 68 | 20.2 |
|  | Piped water | 45 | 13.4 |
|  | Well | 25 | 7.4 |
|  | Piped + River | 24 | 7.1 |
|  | Piped + Pond | 5 | 1.5 |
|  | Well + River | 4 | 1.2 |
|  | Piped + Well | 1 | 0.3 |
| **Contact with stagnant water** | Yes | 309 | 92.0 |
|  | No | 27 | 8.0 |
| **Type of contact** | Washing/fetching water | 253 | 81.9 |
|  | Bathing | 217 | 64.6 |
|  | Walking | 37 | 11.0 |
|  | Washing/fetching +<br>bathing | 28 | 8.3 |
|  | Washing/fetching +<br>walking | 7 | 2.1 |
|  | All three activities | 1 | 0.3 |
| **Frequency of contact** | >2 times/week | 232 | 69.0 |
|  | Seasonally | 55 | 16.4 |
|  | <2 times/week | 22 | 6.5 |
| **Previous UGS infection** | Yes | 57 | 17.0 |
|  | No | 279 | 83.0 |
| **Sought treatment (among infected)** | Yes | 48 | 84.2 |
|  | No | 9 | 15.8 |

### Clinical characteristics of the population

Out of 336 urine samples analysed, the majority were PDT negative (98.2%), with only six samples (1.8%) testing positive. Most samples were nitrite negative (95.5%) and leucocyte negative (89.6%). Among leucocyte-positive samples, 3.0% had moderate (++), and 7.4% had high (+++) counts. In the combined nitrite–leucocyte analysis, nitrite-negative/leucocyte-negative results were predominant (85.7%), followed by nitrite-negative/leucocyte++ (7.1%) and nitrite-positive/leucocyte-negative (3.9%). Haematuria was detected in 4.8% of samples, while schistosome eggs were observed in 13.7% of participants, 10.1% with fewer than 50 eggs/10 mL and 3.6% with 50 or more eggs/10 mL of urine. The distribution of urinalysis and parasitological findings is presented in **Table 3**.

**Table 3.** Clinical and urinalysis characteristics of participants (N = 336)

| Parameter | Characteristics | n | % |
| --- | --- | --- | --- |
| **PDT result** | Negative | 330 | 98.2 |
|  | Positive | 6 | 1.8 |
| **Nitrite** | Negative | 321 | 95.5 |
|  | Positive | 15 | 4.5 |
| **Leucocytes** | Negative | 301 | 89.6 |
|  | ++ | 10 | 3.0 |
|  | +++ | 25 | 7.4 |
| **Combined Nitrite–Leucocyte result** | Nitrite– / Leucocyte– | 288 | 85.7 |
|  | Nitrite– / Leucocyte++ | 24 | 7.1 |
|  | Nitrite– / Leucocyte+ | 9 | 2.7 |
|  | Nitrite+ / Leucocyte– | 13 | 3.9 |
|  | Nitrite+ / Leucocyte++ | 1 | 0.3 |
|  | Nitrite+ / Leucocyte+++ | 1 | 0.3 |
| **Haematuria** | Negative | 320 | 95.2 |
|  | Positive | 16 | 4.8 |
| **Schistosome eggs (per 10 mL urine)** | Negative | 290 | 86.3 |
|  | <50 eggs | 34 | 10.1 |
|  | ≥50 eggs | 12 | 3.6 |

### Prevalence of urogenital schistosomiasis

A total of 336 women of reproductive age were screened for *Schistosoma haematobium* infection using urine microscopy. The overall prevalence of urogenital schistosomiasis was 13.7% (95% CI: 10.2–17.8). Considerable variation was observed across sub-counties, wards, and sub-locations. Prevalence ranged from 2.6% in Sabaki Ward (Magarini Sub-county) to 25.0% in Marikebuni Sub-location (Magarini Ward). Magarini Sub-county recorded a slightly higher prevalence (14.9%) than Rabai (12.0%), with the highest ward-level prevalence found in Magarini Ward (18.8%). Detailed prevalence estimates by administrative unit are presented in **Table 4**.

**Table 4.** Prevalence of urogenital schistosomiasis in women of reproductive age (15 – 50 years) by Sub-county, Ward, and Sub-location (N=336)

| Sub-county | Ward | Sub-location | n Positive | Prevalence (%) | 95% CI |
| --- | --- | --- | --- | --- | --- |
| Rabai | Ruruma | Jimba | 14 | 14.6 | 8.21–23.26 |
|  |  | Mleji | 3 | 6.5 | 1.37–17.90 |
|  | Ward total |  | 17 | 12.0 | 7.13–18.48 |
| Magarini | Magarini | Pumwani | 13 | 15.5 | 8.51–25.01 |
|  |  | Marikebuni | 11 | 25.0 | 13.19–40.34 |
|  | Ward total |  | 24 | 18.8 | 12.40–26.60 |
|  | Garashi | Bore Singwanya,<br>Masindeni, Mikuyuni | 4 | 14.8 | 4.19–33.73 |
|  | Sabaki | Sabaki | 1 | 2.6 | 0.06–13.48 |
|  | Sub-county total |  | 29 | 14.9 | 10.25–20.76 |
| Overall |  |  | 46 | 13.7 | 10.2–17.8 |

### Factors associated with *S. haematobium* in women of reproductive age, Kilifi County Univariate association with *S. haematobium* infection

A univariate model was used to screen for factors associated with *S. haematobium* infection among women of reproductive age in Kilifi County (**Tables 5 and 6).**

**Table 5.**
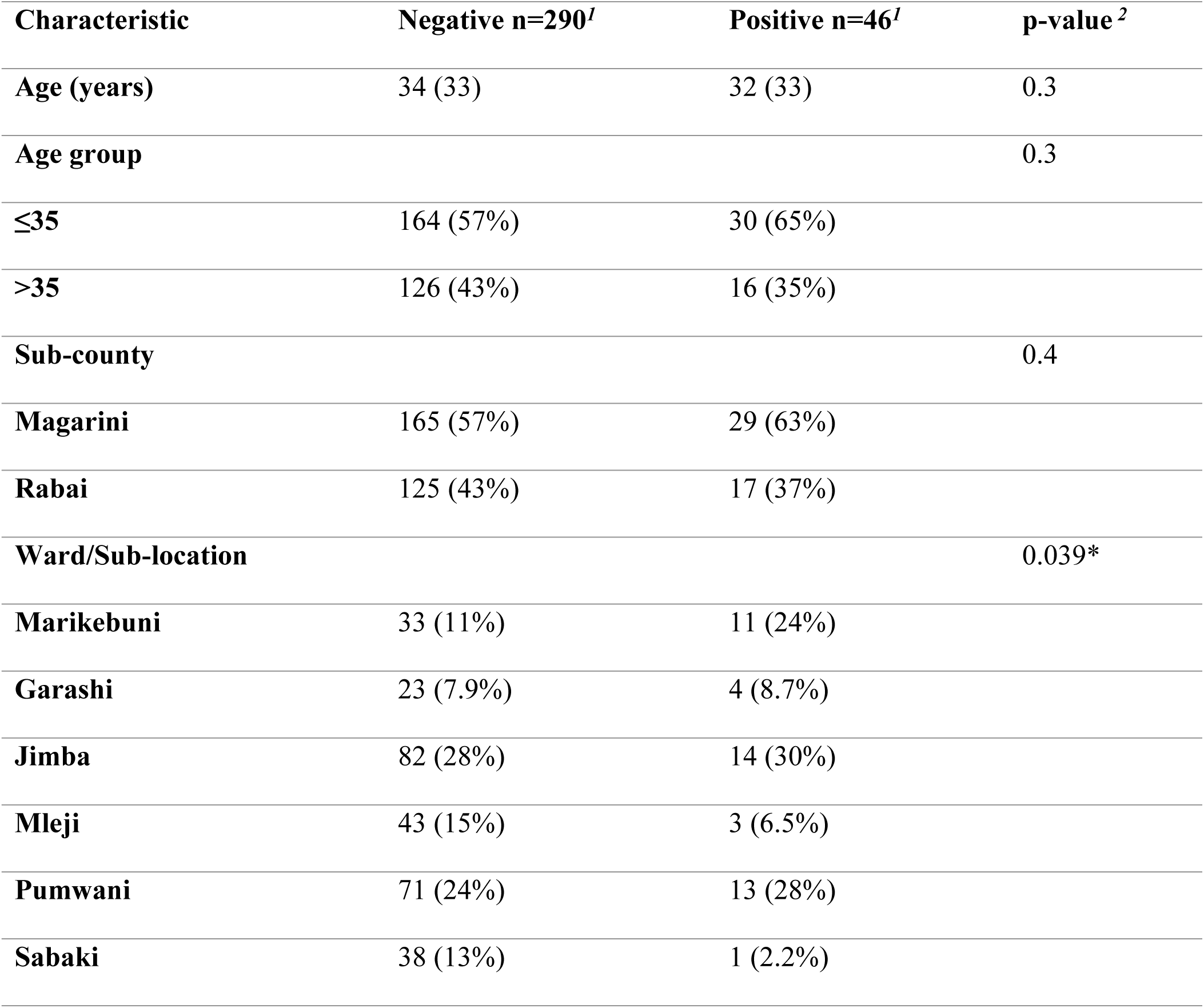
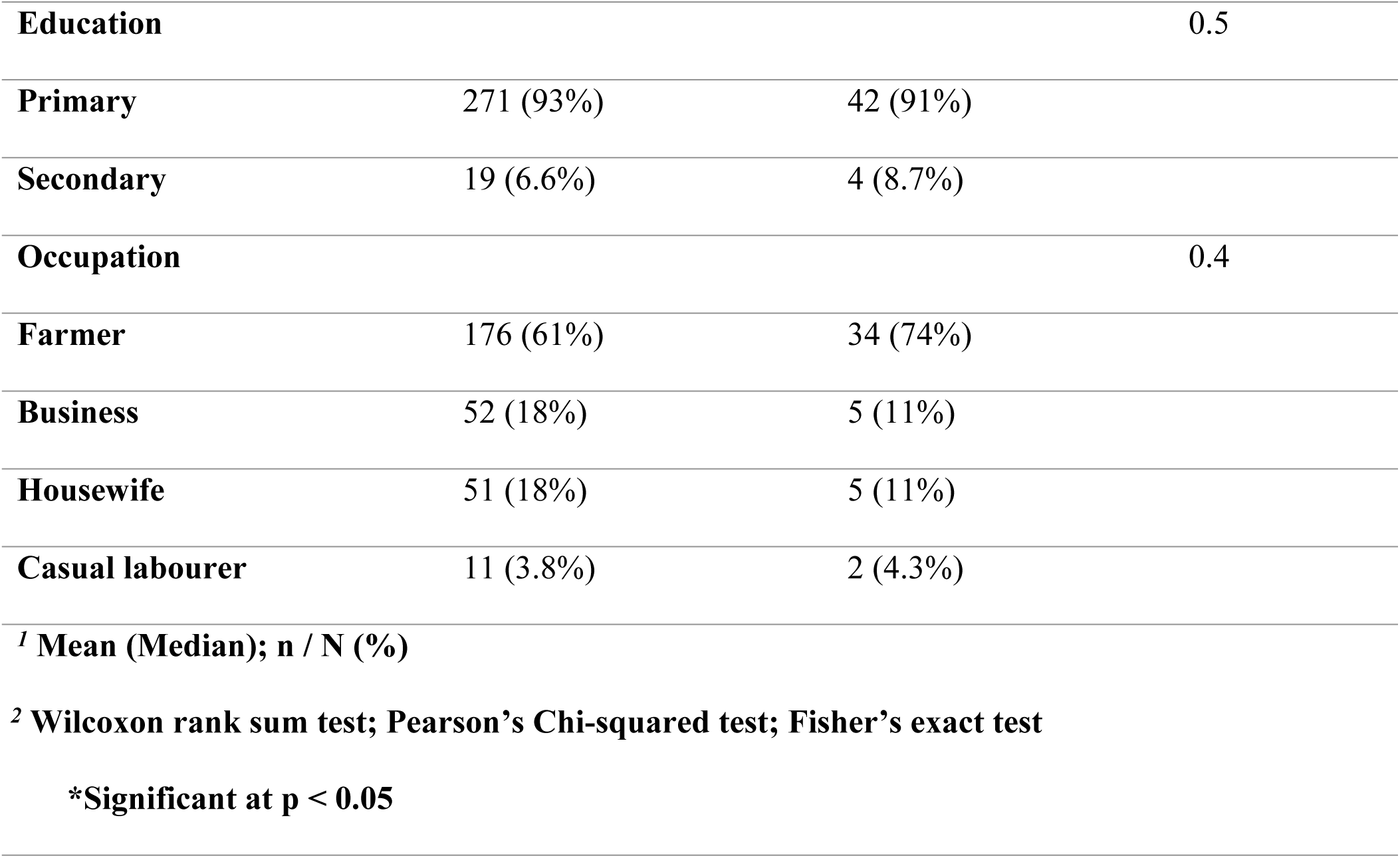
Sociodemographic characteristics associated with *S. haematobium* infection among women of reproductive age in Kilifi County (Rabai and Magarini sub-counties)

**Table 6.**
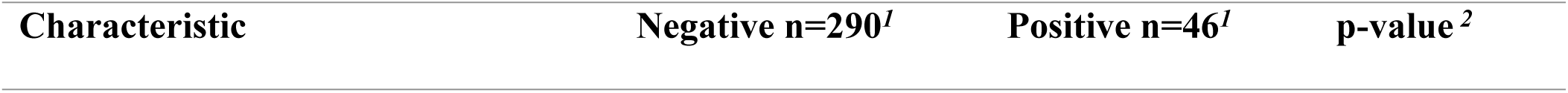
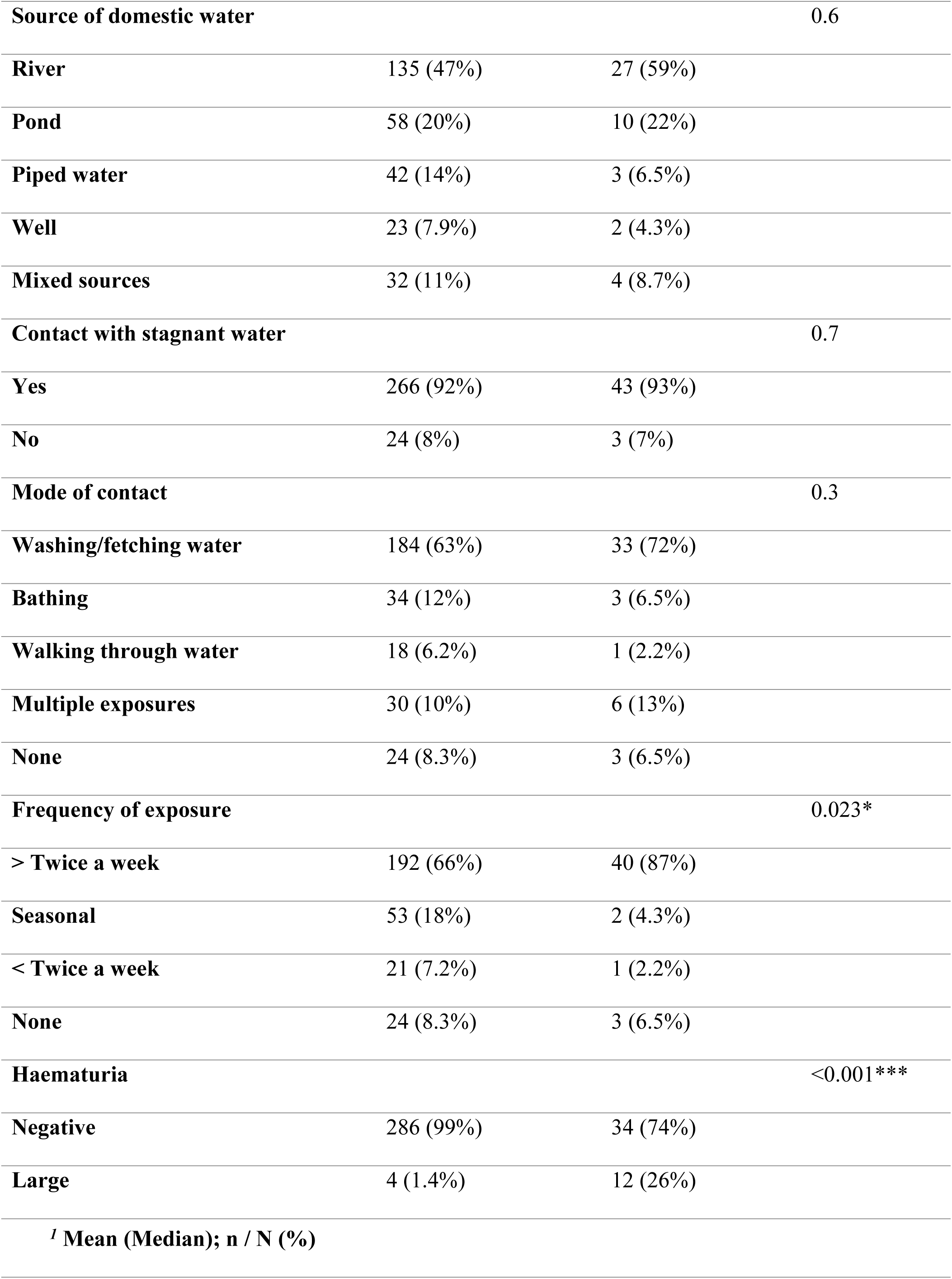
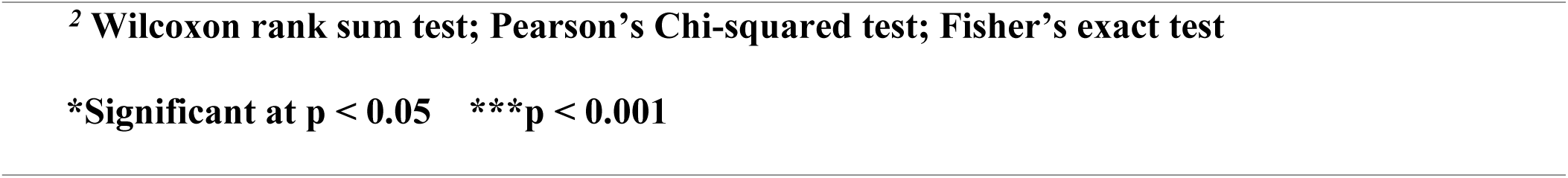
Water contact and clinical characteristics associated with *S. haematobium* infection.

The median age of participants was 33 years (IQR: 14 years). There was no significant association between age group and infection status (p = 0.3) or between sub-county and infection (p = 0.4). Prevalence varied significantly across wards/sub-locations (p = 0.039), with the highest levels observed in Jimba (30%), Pumwani (28%), and Marikebuni (24%), and the lowest in Sabaki (2.2%). (**Table 5**).

No significant associations were found for occupation (p = 0.4), education (p = 0.5), or main source of domestic water (p = 0.6). However, infection was slightly higher among those using river water (59%) compared to other water sources. Contact with stagnant water was common (93%) but not statistically significant (p = 0.7). A significant relationship was noted between frequency of exposure to stagnant water and infection (p = 0.023), with participants exposed more than twice per week showing a higher prevalence (87%). Haematuria was the most strongly associated factor (p < 0.001) (**Table 6).**

### Bivariate association with *S. haematobium* infection

In the bivariate logistic regression (**Table** 7), only haematuria remained a strong predictor of infection. Participants with visible haematuria were 25.24 times more likely to be infected (*OR* = 25.24, 95% CI: 7.07–82.63, p < 0.001). No statistically significant associations were observed for age group, sub-county, education level, or reported contact with stagnant water.

**Table 7:** Bivariate logistic regression of factors associated with *S. haematobium* infection among women of reproductive age, Kilifi County (Rabai and Magarini sub-counties)

| Characteristic | Negative<br>(n=290) | Positive<br>(n=46) | OR (95% CI) | P-value |
| --- | --- | --- | --- | --- |
| **Age group (years)** |  |  |  |  |
| ≤35 | 164 (56.6%) | 30 (65%) | 1.4 (0.75–2.76) | 0.27 |
| >35 | 126 (43.4%) | 16 (35%) | Ref |  |
| **Sub-county** |  |  |  |  |
| Magarini | 165 (57%) | 29 (63%) | 1.3 (0.68–2.46) | 0.43 |
| Rabai | 125 (43%) | 17 (37%) | Ref |  |
| **Haematuria** |  |  |  |  |
| Large | 4 (1.4%) | 12 (26.1%) | 25.24 (7.07–82.63) | <0.001*** |
| Negative | 286 (98.6%) | 34 (73.9%) | Ref |  |
| **Education level** |  |  |  |  |
| Primary | 271 (93.4%) | 42 (91.3%) | 1.4 (0.44–4.9) | 0.6 |
| Secondary | 19 (6.6%) | 4 (8.7%) | Ref |  |
| **Contact with stagnant water** |  |  |  |  |
| Yes | 266 (92%) | 43 (93%) | 1.3 (0.4–4.5) | 0.69 |
| No | 24 (8%) | 3 (7%) | Ref |  |
| ***p < 0.001; OR = Odds Ratio; CI = Confidence Interval; Ref = Reference category |  |  |  |  |

### Factors associated with *S. haematobium* infection

Binomial logistic regression was conducted to determine factors independently associated with *S. haematobium* infection. The predictors included location, occupation/economic activity, source of domestic water, mode of contact, and frequency of contact with stagnant water. The sub-county of residence was not significantly associated with infection; however, variations were observed across specific locations. Compared to Sabaki (reference category), participants residing in Marikebuni had significantly higher odds of infection (OR = 12.67; 95% CI: 1.55–103.39; *p* = 0.02). Jimba (OR = 6.45; 95% CI: 0.82–51.15; *p* = 0.08), Pumwani (OR = 6.96; 95% CI: 0.88–55.24; *p* = 0.07), and Garashi (OR = 6.61; 95% CI: 0.70–62.81; *p* = 0.10) also had higher odds, although these associations were not statistically significant. Regarding occupation, farmers had the highest prevalence (73.9%) and were twice as likely to be infected compared to businesswomen (reference group), although the difference was not statistically significant (OR = 2.00; 95% CI: 0.75–5.40; *p* = 0.17). Similarly, housewives (OR = 1.02; 95% CI: 0.28–3.74; *p* = 0.98) and casual labourers (OR = 1.90; 95% CI: 0.32–11.04; *p* = 0.48) did not differ significantly from the reference group.

Participants using river water for domestic purposes had the highest infection prevalence (59%); however, this association did not reach statistical significance (OR = 2.80; 95% CI: 0.81–9.70; *p* = 0.10). Exposure behaviours also influenced the likelihood of infection. Those who washed or fetched water from stagnant sources had greater odds of infection (OR = 1.43; 95% CI: 0.41–5.04; *p* = 0.57) compared to those without such contact. Participants reporting contact more than twice weekly had higher infection odds (OR = 1.67; 95% CI: 0.48–5.80; *p* = 0.42) compared to those reporting seasonal or no exposure. However, none of these associations reached statistical significance (**Table 8**).

**Table 8.**
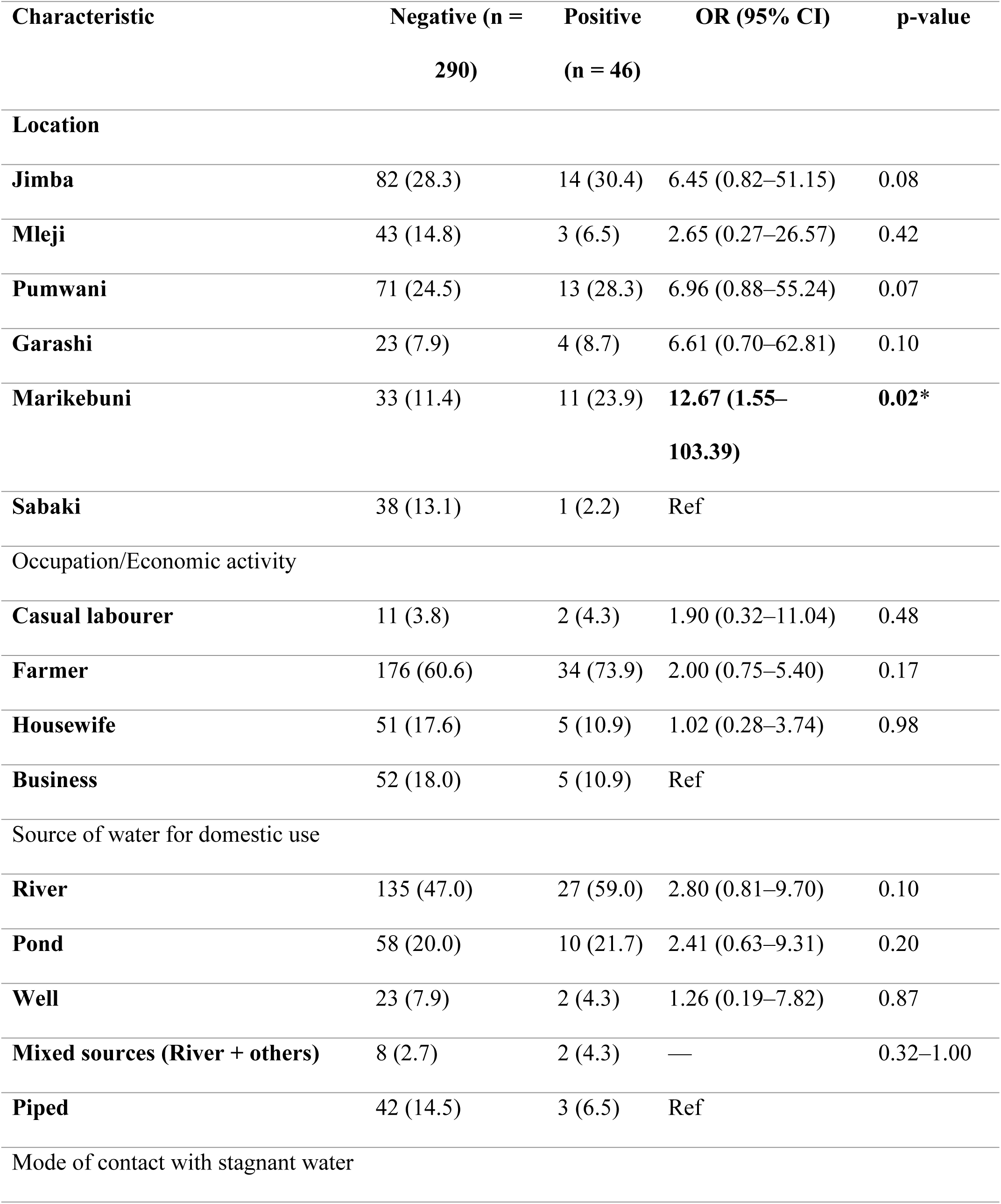
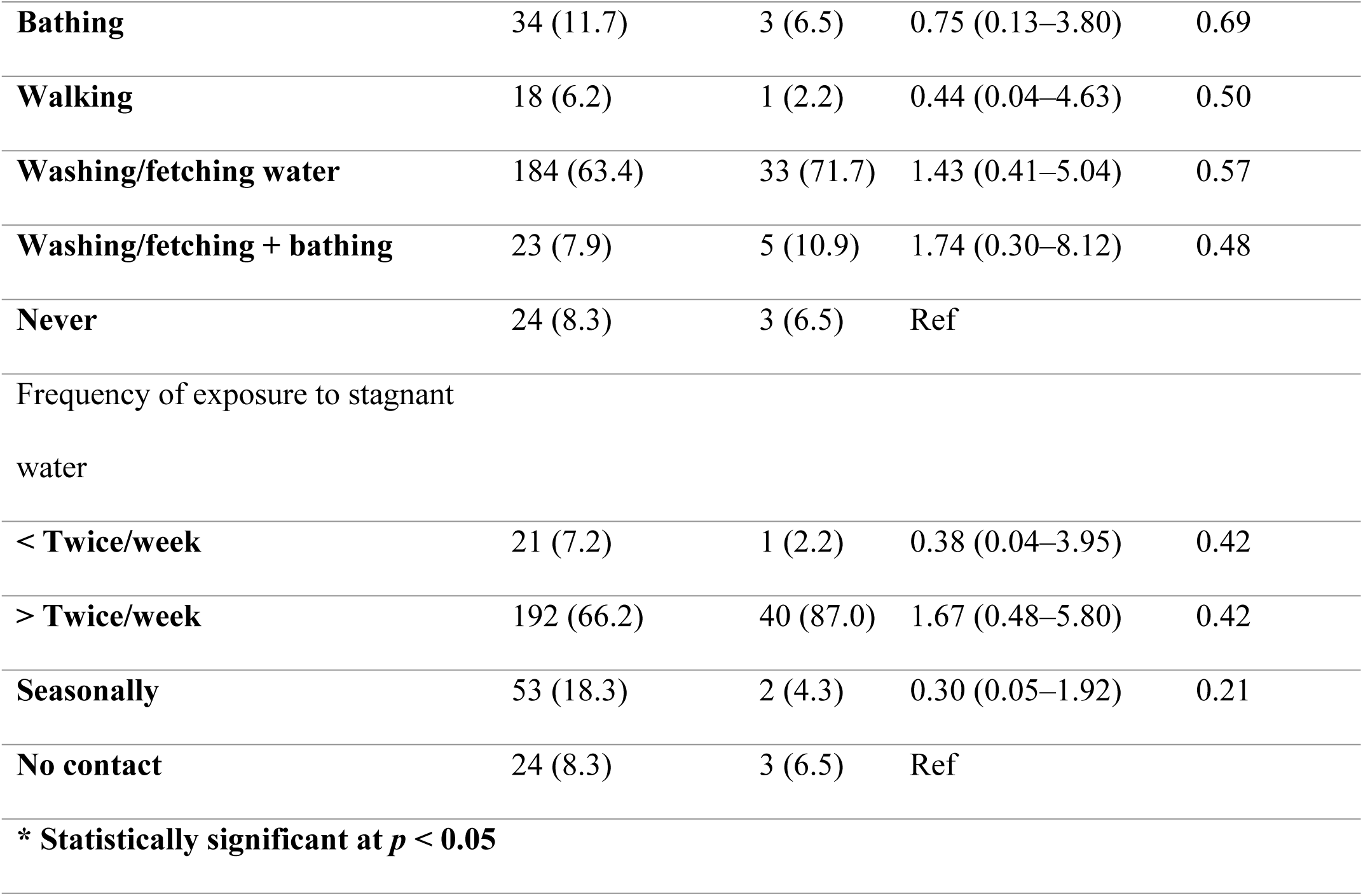
Factors Associated with *S. haematobium* infection among women of reproductive age, Kilifi County.

| Characteristic | Negative (n =<br>290) | Positive<br>(n = 46) | OR (95% CI) | p-value |
| --- | --- | --- | --- | --- |
| **Location** |  |  |  |  |
| **Jimba** | 82 (28.3) | 14 (30.4) | 6.45 (0.82–51.15) | 0.08 |
| **Mleji** | 43 (14.8) | 3 (6.5) | 2.65 (0.27–26.57) | 0.42 |
| **Pumwani** | 71 (24.5) | 13 (28.3) | 6.96 (0.88–55.24) | 0.07 |
| **Garashi** | 23 (7.9) | 4 (8.7) | 6.61 (0.70–62.81) | 0.10 |
| **Marikebuni** | 33 (11.4) | 11 (23.9) | **12.67 (1.55–**<br>**103.39)** | **0.02\*** |
| **Sabaki** | 38 (13.1) | 1 (2.2) | Ref |  |
| Occupation/Economic activity |  |  |  |  |
| **Casual labourer** | 11 (3.8) | 2 (4.3) | 1.90 (0.32–11.04) | 0.48 |
| **Farmer** | 176 (60.6) | 34 (73.9) | 2.00 (0.75–5.40) | 0.17 |
| **Housewife** | 51 (17.6) | 5 (10.9) | 1.02 (0.28–3.74) | 0.98 |
| **Business** | 52 (18.0) | 5 (10.9) | Ref |  |
| Source of water for domestic use |  |  |  |  |
| **River** | 135 (47.0) | 27 (59.0) | 2.80 (0.81–9.70) | 0.10 |
| **Pond** | 58 (20.0) | 10 (21.7) | 2.41 (0.63–9.31) | 0.20 |
| **Well** | 23 (7.9) | 2 (4.3) | 1.26 (0.19–7.82) | 0.87 |
| **Mixed sources (River + others)** | 8 (2.7) | 2 (4.3) | — | 0.32–1.00 |
| **Piped** | 42 (14.5) | 3 (6.5) | Ref |  |
| Mode of contact with stagnant water |  |  |  |  |
| **Bathing** | 34 (11.7) | 3 (6.5) | 0.75 (0.13–3.80) | 0.69 |
| **Walking** | 18 (6.2) | 1 (2.2) | 0.44 (0.04–4.63) | 0.50 |
| **Washing/fetching water** | 184 (63.4) | 33 (71.7) | 1.43 (0.41–5.04) | 0.57 |
| **Washing/fetching + bathing** | 23 (7.9) | 5 (10.9) | 1.74 (0.30–8.12) | 0.48 |
| **Never** | 24 (8.3) | 3 (6.5) | Ref |  |
| Frequency of exposure to stagnant water |  |  |  |  |
| **< Twice/week** | 21 (7.2) | 1 (2.2) | 0.38 (0.04–3.95) | 0.42 |
| **> Twice/week** | 192 (66.2) | 40 (87.0) | 1.67 (0.48–5.80) | 0.42 |
| **Seasonally** | 53 (18.3) | 2 (4.3) | 0.30 (0.05–1.92) | 0.21 |
| **No contact** | 24 (8.3) | 3 (6.5) | Ref |  |
| **\* Statistically significant at $p < 0.05$** |  |  |  |  |

### Independent predictors of *S. haematobium* infection

Multivariate logistic regression analysis was conducted to identify independent predictors of *S. haematobium* infection among women of reproductive age. Variables with a *p* ≤ 0.25 in the univariate analysis were included in the final model. Table 9 presents the adjusted odds ratios (AORs), 95% confidence intervals (CIs), and corresponding *p*-values. Participants aged ≤ 35 years had 1.21 times higher odds of infection compared to those aged > 35 years, although this association was not statistically significant (95% CI: 0.60 – 2.43, p = 0.59). Participants with hematuria were 20.8 times more likely to have *S. haematobium* infection, a statistically significant finding (95% CI: 5.45 – 79.57, p < 0.001). Compared to businesswomen, farmers were 1.65 times more likely to be infected, although this relationship was not significant (95% CI: 0.54 – 5.05, p = 0.38).

**Table 9:** Factors associated with *S. haematobium* (P≤0.25)

| Parameter | B(SE) | OR | 95% CI | P-value |
| --- | --- | --- | --- | --- |
| **Age** |  |  |  |  |
| ≤35 | 0.19 (0.33) | 1.21 | (0.60 – 2.43) | 0.59 |
| **Location** |  |  |  |  |
| Jimba | 1.42 (1.10) | 4.12 | (0.45 – 37.59) | 0.21 |
| Mleji | 0.60 (1.23) | 1.82 | (0.17 – 19.15) | 0.62 |
| Pumwani | 1.47 (1.12) | 4.35 | (0.48 – 39.40) | 0.19 |
| Garashi | 1.36 (1.20) | 3.9 | (0.38 – 39.72) | 0.286 |
| Marikebuni | 2.19 (1.11) | 8.95 | (1.01 – 79.28) | 0.049* |
| Sabaki | Ref |  |  |  |
| **Hematuria** |  |  |  |  |
| Positive | 3.04 (0.73) | 20.83 | (5.45 – 79.57) | <0.001*** |
| **Occupation** |  |  |  |  |
| Casual Laborer | 0.22 (1.01) | 1.25 | (0.18 – 8.79) | 0.82 |
| Farmer | 0.5(0.57) | 1.65 | (0.54 - 5.05) | 0.38 |
| Housewife | -0.09 (0.77) | 0.91 | (0.21 – 3.99) | 0.9 |
| Business | Ref |  |  |  |
| **Frequency of contact with water** |  |  |  |  |
| Less than twice a week | -1.14 (1.18) | 0.32 | (0.03– 53.37) | 0.34 |
| More than twice a week | 0.34 (0.67) | 1.41 | (0.37 – 5.38 | 0.61 |
| Seasonally | -1.27 (1.03) | 0.28 | (0.04 – 2.01) | 0.21 |
| Never | Ref |  |  |  |
| **Constant** | **-3.77 (1.35)** | **-1.34** |  | **<0.001** |

**p < 0.05; \*\* p < 0.01; \*\*\* p < 0.001**
**AOR = Adjusted Odds Ratio; CI = Confidence Interval; B= beta; SE = Standard Error**

In terms of location, participants from Marikebuni had significantly higher odds of infection compared to those from Sabaki (AOR = 8.95, 95% CI: 1.01 – 79.28, p = 0.049). Other locations, Jimba, Pumwani, Mleji, and Garashi, showed elevated odds but were not statistically significant. Participants who reported contact with water more than twice a week were 1.41 times more likely to be infected than those who did not, although the difference was not significant (95% CI: 0.37 – 5.38, p = 0.61).

## Discussion

The study employed a two-phase approach. Urine samples were collected over three consecutive days in line with WHO recommendations for *S. haematobium* diagnosis, accounting for day-to-day variation in egg excretion and enhancing diagnostic sensitivity. Subsequently, questionnaires were administered and genital samples were collected under the oversight of the principal investigator, ensuring correct participant enrolment and enhancing the validity of both parasitological and self-reported data.

A significant proportion of participants were aged 35 years or younger, reflecting the demographic structure of women of reproductive age in the study area. Since recruitment was carried out systematically at the household level and before any diagnostic testing, the observed age distribution aligns with the underlying population rather than disease-related selection. The higher representation from Magarini sub-county (63.4%) corresponds with its larger population and the proportional allocation used during sampling. Sub-locations such as Marikebuni and Pumwani contributed more participants due to the greater number of eligible households in these areas. The dominance of farming (62.5%) as the main occupation is characteristic of rural Kilifi County, where agriculture is the primary livelihood. These occupational patterns also indicate routine contact with natural water sources, which may increase exposure to schistosomiasis. Overall, the demographic and geographic features of the sample reflect the population structure and livelihood context of the study area, offering a consistent basis for interpreting the observed patterns of schistosomiasis exposure.

Reliance on river water (48.2%) for domestic use, alongside substantial use of pond water (20.2%) and limited access to piped water (13.34%), indicates regular interaction with natural water bodies where *S. haematobium* cercariae may be present. Almost all participants (92%) reported frequent contact with stagnant water, and a large proportion engaged in water-related tasks such as fetching and washing (81.88%), highlighting routine behaviours that increase exposure opportunities. Although exposure duration was not quantified, the observation that 69% of participants encountered stagnant water more than twice per week suggests repeated and likely sustained interaction with potentially contaminated sources, reinforcing the environmental conditions that facilitate transmission.

Low levels of educational attainment were also notable, with 93.2% of participants having completed only primary education or less. Such limited educational exposure may influence health literacy, awareness of schistosomiasis risk factors, and uptake of preventive behaviours. (26). Despite 17% reporting a previous history of schistosomiasis, only 15.8% had ever sought treatment, highlighting persistent gaps in healthcare access, disease recognition, and knowledge of available services. Collectively, these socio-demographic and environmental characteristics underscore a combination of behavioural, structural, and ecological factors that shape exposure patterns and may contribute to ongoing transmission within the study setting

### Prevalence and Geographic Patterns

The overall prevalence of urogenital schistosomiasis (UGS) in Kilifi County was 13.7%, with Magarini Sub-county being slightly higher (14.9%) than Rabai (12%), differences that may reflect contrasting ecological and infrastructural characteristics. Magarini is traversed by permanent freshwater sources such as the Sabaki River, which provide suitable habitats for *Bulinus* spp. snails, the intermediate hosts for *S. haematobium*. Additionally, the sub-county is relatively more marginalised, with lower government service penetration compared to Rabai, potentially affecting the reach of schistosomiasis prevention and control efforts (27).

The prevalence observed in this study (13.7%) is lower than that reported in a previous study, which recorded 22.3% (14). Although both studies sampled from Magarini and Rabai Sub-counties, the earlier study included only Boyani, Jimba, and Kanyumbuni sub-locations in Rabai, and two sub-locations in Magarini (14). In contrast, our survey covered **f**our wards in Magarini and two sub-locations in Rabai, providing broader geographic coverage. Therefore, we believe that we have captured a more representative cross-section of the population, including areas with lower transmission intensity, which could account for the lower overall prevalence we observed. A study in neighbouring Kwale County reported a significantly lower prevalence of 3.8% among women of reproductive age (28). These variations likely reflect differences in environmental exposure, proximity to freshwater bodies, local transmission intensity, the effectiveness of ongoing control interventions, and population-level practices such as sanitation, access to preventive chemotherapy, and community awareness.

### Spatial Heterogeneity and Local Transmission Zones

Residence was significantly associated with UGS status. In Rabai Sub-county, Jimba sub-location within Ruruma ward had the highest prevalenc**e (**14.6%), indicating a persistent transmission hotspot. In Magarini ward, Marikebuni recorded the highest prevalence at 25%, followed by Pumwani at 15.5%. These patterns suggest micro-geographic clustering of transmission and underscore the need for intensified, location-specific interventions focusing on improved sanitation, health education, and access to treatment.

### Age, Water Contact, and Behavioural Risk Factors

Although UGS prevalence was higher among younger women, the difference was not statistically significant. This trend aligns with findings from other studies reporting greater infection rates among younger age groups, who typically engage more in water-related activities such as swimming, fetching water, and bathing (29–32). Participants reporting frequent contact with stagnant water during domestic chores were also more likely to be infected, reinforcing the role of routine water-contact behaviours in exposure to cercariae. *Bulinus* snails thrive in stagnant or slow-flowing freshwater, which explains why infection rates were higher among individuals reporting frequent exposure to such environments (4,33).

Women engaged in farming were approximately twice as likely to have UGS compared to housewives and casual labourers. This association is consistent with previous studies demonstrating farming as a major risk factor due to regular exposure to contaminated water during agricultural activities such as irrigation (34–37). However, contrasting results from Malawi found no association between economic activity and *S. haematobium* infection (29), highlighting how local ecological and behavioural contexts influence risk.

### Socioeconomic and Educational Influences

Schistosomiasis is well recognised as a disease of poverty, disproportionately affecting communities with low literacy, poor sanitation, and limited access to clean water and healthcare (38). In the present study, UGS prevalence was higher among women with lower education levels, which may reflect restricted access to health information, preventive practices, and timely treatment. The reliance on river water for domestic activities, associated with the highest infection prevalence, further indicates that environmental and infrastructural limitations are present.

### Health-Seeking Behaviour and Treatment Gaps

Health-seeking behaviour among participants was generally poor, with low proportions reporting having sought treatment despite previous schistosomiasis infections. This suggests barriers such as limited access to healthcare facilities, low awareness of disease symptoms, cultural perceptions, and stigma. In contrast, a study in neighbouring Kwale County reported generally positive health-seeking behaviour, although concerns related to shame and genital examinations were noted as barriers to testing (18).

### Co-Infections, Differential Diagnosis, and Diagnostic Implications

Urinary tract infections (UTIs) can complicate the clinical diagnosis of UGS, as symptoms such as dysuria overlap with those of *S. haematobium* infection (7,39). Dipstick urinalysis provides a practical point-of-care tool in low-resource settings. In the present study, a small number of participants tested positive for nitrites (15/336), which are typically associated with UTIs, dysuria, and sexually transmitted infections (STIs). These inflammatory markers may therefore reflect co-existing UTIs rather than schistosomiasis, while overlapping nitrite and leukocyte findings in approximately 1% of participants may suggest a possible STI co-infection (40).

### Haematuria as a Diagnostic Indicator

Haematuria, a hallmark of urogenital schistosomiasis (UGS), results from *Schistosoma haematobium* eggs penetrating the bladder wall and ureters. As the eggs migrate, they cause mechanical disruption of small blood vessels and trigger inflammatory responses, leading to micro- and macrohaemorrhages in the urinary tract (41–43). In the present study, haematuria was a strong predictor of infection: participants with haematuria were 20.83 times more likely to have UGS compared to those without, even after adjusting for potential confounders. This reinforces haematuria as a reliable clinical indicator in endemic settings, particularly where laboratory diagnostics may be limited. Integrating urine dipstick tests or other rapid, point-of-care screening methods into routine surveillance could enhance early detection and timely treatment, reducing long-term complications such as bladder fibrosis or ureteral obstruction (43). Despite this strong association, haematuria was not present in all infected individuals, highlighting the need for a combined diagnostic approach that incorporates both symptom-based screening and parasitological confirmation through urine microscopy or molecular techniques. Health education campaigns should emphasise recognition of haematuria and other clinical signs of UGS to encourage prompt healthcare-seeking behaviour and improve early diagnosis and treatment.

### Study limitations

Participation in the second phase, which involved the administration of questionnaires and the collection of genital samples, was lower than in the first phase. Women who were menstruating or had engaged in sexual activity within 48 hours of the second phase were excluded to maintain th**e** integrity of the subsequent diagnostic results, which in this phase primarily pertained to questionnaire responses. Additional barriers, including cultural norms, logistical challenges, and personal constraints, further reduced completion rates for these components, potentially limiting the generalisability of the risk factor analysis. Self-reported behaviours may also be subject to recall or social desirability bias. Nonetheless, the overall sample size for prevalence estimation was achieved, allowing a robust assessment of UGS prevalence. The two-phase design still provided a comprehensive evaluation of both prevalence and associated risk factors while maintaining ethical and methodological rigour.

## Conclusion

The overall prevalence of urogenital schistosomiasis among women of reproductive age in Kilifi County was 13.7%, with Magarini sub-county showing a higher prevalence than Rabai, largely due to the presence of the Sabaki River, a key source of water and livelihood. This underscores the need for strengthened water, sanitation, and hygiene interventions to reduce the disease burden. Haematuria, while long recognised as an indicator of *S. haematobium* infection, demonstrated particular utility among women in endemic settings, serving as a simple, non-invasive, and reliable marker. Its integration into community-level screening and point-of-care programmes can facilitate early detection and prompt treatment. Furthermore, haematuria-based screening can support symptom-driven health education campaigns, encouraging timely healthcare-seeking among at-risk women and contributing to more effective schistosomiasis control in these communities.

## Data Availability

All relevant data supporting the findings of this study are provided within the manuscript and its Supporting Information files.

## Acknowledgments

We sincerely thank all the women who participated in this study for their time and willingness to contribute. We are grateful to the Kilifi County Department of Health and the sub-county health management teams for their support and collaboration during field activities. We acknowledge the assistance of local health facility staff and community health volunteers who facilitated participant mobilisation and sample collection. We also thank the laboratory teams for their technical support in sample processing and analysis.

The first author acknowledges support from the Organisation for Women in Science for the Developing World (OWSD) through the Sandwich Programme, which assisted her research training. This support did not influence the study design, data collection, analysis, or interpretation. Finally, we appreciate the guidance and mentorship provided by our supervisors throughout the study.

